# Assessing the Role of Patient Generation Techniques in Virtual Clinical Trial Outcomes

**DOI:** 10.1101/2024.06.11.24308775

**Authors:** Jana L. Gevertz, Joanna R. Wares

**Author notes:** Contributing authors.

## Abstract

Virtual clinical trials (VCTs) are growing in popularity as a tool for quantitatively predicting heterogeneous treatment responses across a population. In the context of a VCT, a plausible patient is an instance of a mathematical model with parameter (or attribute) values chosen to reflect features of the disease and response to treatment for that particular patient. A number of techniques have been introduced to determine the set of model parametrizations to include in a virtual patient cohort. These methodologies generally start with a prior distribution for each model parameter and utilize some criteria to determine whether a parameter set sampled from the priors should be included or excluded from the plausible population. No standard technique exists, however, for generating these prior distributions and choosing the inclusion/exclusion criteria. In this work, we rigorously quantify the impact that VCT design choices have on VCT predictions. Rather than use real data and a complex mathematical model, a spatial model of radiotherapy is used to generate simulated patient data and the mathematical model used to describe the patient data is a two-parameter ordinary differential equations model. This controlled setup allows us to isolate the impact of both the prior distribution and the inclusion/exclusion criteria on both the heterogeneity of plausible populations and on predicted treatment response. We find that the prior distribution, rather than the inclusion/exclusion criteria, has a larger impact on the heterogeneity of the plausible population. Yet, the percent of treatment responders in the plausible population was more sensitive to the inclusion/exclusion criteria utilized. This foundational understanding of the role of virtual clinical trial design should help inform the development of future VCTs that use more complex models and real data.

## 1 Introduction

Clinical trials are research studies where novel medical interventions are tested on people who volunteer to receive the treatment. These studies are the primary way that researchers find out if a new treatment is safe and effective in humans. Clinical trials for new drug therapies have four phases, with the number of needed participants increasing for each phase. By phase four of a clinical trial, thousands of participants who have the condition or disease that the novel therapeutic can possibly treat are required for the study [1].

There has been an increased focus on ensuring that clinical trials include subjects that are as diverse as those that are affected by the particular condition or disease. The Food and Drug Administration has been working to create initiatives and policies to increase diversity in clinical trials [2], including guidance for researchers on how to create diversity plans to improve enrollment of participants from underrepresented racial and ethnic populations [3].

But as they state, “Increasing representation is a multi-faceted challenge that will require collaboration of our federal partners, industry, health care professionals, patient advocacy groups and community-based organizations” [3]. There are barriers to entry for participation that are specific to racial and ethnic groups that can make achieving equitable representation difficult [3]. Achieving a proper gender balance has also been a neglected feature of many clinical trials, although the situation appears to be somewhat improving in this regard [4]. For some conditions women represent upward of 60% of those afflicted but are still barely 40% of the subjects in clinical trials [4]. The predictions made by clinical trials are thus limited by small sample sizes and may be biased to certain demographic groups.

Virtual clinical trials (VCTs) are growing in popularity as a tool for predicting and quantifying the uncertainty of the effects of therapy on disease progression [5, 6]. VCTs combine data-validated mathematical models with computational techniques to enhance the efficiency, increase the success rates, and decrease the costs associated with the drug development process [7]. In this manuscript we adopt the VCT terminology used by Rieger and colleagues [8]. First, a plausible patient (PP) is a parametrization of the model that is deemed biologically feasible based on defined parameter and model output constraints. In other words, a PP should be thought of as an instance of a mathematical model with parameter (or attribute) values chosen to reflect a feature of the disease for that particular patient. An ensemble of these biologically reasonable plausible patients that could be included in a clinical trial is called a plausible population (PPop). If data is available to further determine how likely a PP is to be in a clinical trial, that data is utilized to identify a subset of PPs called virtual patients (VPs). This collection of VPs is called a virtual population (Vpop) [8]. VCTs have been utilized across a number of medical conditions, including cancer [9–13], infectious diseases [14, 15], cardiovascular disease [16–18], rheumatoid arthritis [19], and diabetes [17, 18, 20]. These studies and others demonstrate the value of VCTs for rational protocol design that accounts for inter-patient heterogeneity.

A recent review paper [21] has introduced a step-by-step best practice guide for conducting a model-based virtual clinical trial. The process that the authors’ outlined can be cyclical, where knowledge gained at one step may necessitate returning to an earlier step. Though, we will present the steps in linear order here. A VCT begins with defining the question of interest for the study; the authors call this VCT Step 0 [21]. For instance, one may be interested in quantifying how the percent of responders changes as a function of drug dose (see Figure 3 in Craig et al. [21]). Other potential questions of interest include the impact of the number of doses, the spacing between doses, combining therapies, etc. Given a question of interest, the next step of the VCT process is to create a data-informed model that is complex enough to capture the impacts of the drug, but not too complicated that parametrizing and interpreting the model becomes intractable (Step 1). Best practices should be used to parameterize the model (Step 2), for instance by fitting model parameters to a training dataset and saving separate data for validation purposes [21]. As an example, in a preclinical setting, the data for informing the model in Step 1 could come from murine tumor growth experiments. Even under ideal experimental conditions, sample sizes for this step are typically very small, with 10 mice or fewer per condition being common.

Following model parametrization, it is important to further understand the structure of parameter space (Step 3), which can be done using sensitivity and identifiability analyses [21]. This step is especially important for identifying the specific parameters (patient characteristics) that will be used to define a virtual patient. Selecting too few parameters at this step (and thus fixing too many parameters) may result in a cohort of virtual patients that is not sufficiently heterogeneous, whereas selecting too many parameters to vary across virtual patients greatly increases the computational complexity of conducting a virtual clinical trial. Further, there are often model parameters that simply do not vary much from patient-to-patient, and there are often parameters that the model is highly insensitive to. Fixing these parameters in the model, rather than allowing them to vary across virtual patients, is thus a reasonable choice.

Once the parameter set that will vary across virtual patients is defined, plausible patients can be created (Step 4). For instance, if the disease of interest were some type of cancer, then a PP could be a differential equation with parameters that represent the intrinsic growth rate of a patient’s tumor and their response to a specific therapeutic. In theory, the parametrization of a plausible patient requires knowing the distribution of the parameter in the population from preclinical or clinical data. Given the general unavailability of this information, a number of techniques have been introduced to generate PPs for a VCT. Since there is no standardized technique for creating a plausible patient, this raises the following question: does the method used to create the plausible patient population bias the results of the trial, as occurs in real-world clinical trials?

In this study, we investigate whether the method chosen to create a virtual patient population impacts the variability in the PPops themselves (that is, in the parameters “included” in the virtual clinical trial), and/or the predictions about treatment response (that is, the answer to the question posed in the VCT). This work is similar in spirit to [22] which sought to compare four different approaches to generate VPops. The studies differ in several key ways, however. In Kolesova et al. [22], they sought to create VPops of the same size as the experimental data, whereas we are interested in using PPs to create larger samples to better understand the variability found in the population. Further, the parameter inclusion methods in Kolesova et al. were assessed in terms of their ability to generate VPs that are statistically similar to the experimental data. As will be detailed in the Methods, for our analysis, similarity to the experimental data is an inclusion criterion for VPs, not an assessment criteria. Finally, while Kolesova et al. [22] applied several methods to a data-motivated quantitative systems pharmacology model of erythropoiesis in which a VP was defined by 39 model parameters, we intentionally use simulated data and a toy model to isolate the impact of VCT design.

This paper is organized as follows. In Section 2 we describe two commonly-used methods for generating plausible patient populations: the “accept-or-reject” and the “accept-or-perturb” methods. To explore the properties of these methods in a controlled setting, we generate synthetic data from a previously-developed cellular automaton model of tumor growth and radiotherapy [23] and propose a simple ordinary differential equation (ODE) model of the synthetic data. The model used to generate the synthetic data, the ODE model, and the fits of the ODE model to the synthetic data are also described in Section 2. In Section 3, we explore if, and how, virtual clinical trial predictions using plausible populations depend on the method used to generate the plausible patients. In particular, we explore if the choice of the prior parameter distribution impacts the predicted posterior distribution in the PPop within the framework of one method (that is, using either accept-or-reject or accept-or-perturb). We also explore the impact that the method itself has on the posterior distribution in the PPop, and we quantify how predictions made for virtual clinical trials vary across methods. We conclude with a discussion of the implications for incorporating virtual clinical trials into the workflow of drug development.

## 2 Methods

Assessing the role of virtual clinical trial design is complicated by the messiness of real data and the complexity of real models. Herein, we intentionally simplify the patient data and the model used to analyze the data so that we can focus on how various aspects of virtual clinical trial design influence the pool of plausible patients and the prediction of the virtual clinical trial.

For any VCT, the first step (Step 0 in [21]) is defining the question of interest. For our virtual clinical trial, the question of interest will be to determine the efficacy of a fixed dose and schedule of radiotherapy treatment on reducing tumor growth across a heterogeneous patient population. In this Section, we first detail how we generated a simulated patient dataset that forms the foundation of our VCT. Next, we present and parametrize a toy “data-informed model” of the simulated patient data (corresponding to Steps 1 and 2 outlined above). The last subsection details two standard methods for generating plausible patients.

### 2.1 Synthetic Patient Data

Any virtual clinical trial requires preclinical or clinical data that is used to inform model development and parametrization. Rather than using data from a clinical trial or an in vivo experiment, which is inherently noisy, we instead generated data from an open source two-dimensional cellular automaton model of spatially-resolved tumor growth treated with radiotherapy [23]. In this model, each automaton element is a two-dimensional cross-section of a three-dimensional tumor spheroid that can either be empty or in one of three cancerous states: proliferating (*P*), quiescent (*Q*), and necrotic (*N*). The state of a cell is determined by the local oxygen concentration. In response to treatment, each non-necrotic cancer cell can be killed by a radiotherapy dose *d* with probability *p_RAD_* = 1 *− e^−αd−βd^*, where *α* and *β* represent radiosensitivity parameters [23].

To generate simulated data for our study, all treatment-related parameters are fixed as specified in [23]. We consider only one cell type, neutral cell-cell interactions, low radiosensitivity (*α/β* = 9, *α* = 0.14), and a low necrotic decay rate (*p_NR_*= 0.004). Tumor growth is initiated by cells occupying 0.25% of the total volume of the available space, and tumors grow for 15 days prior to the start of treatment. Treatment is administered at each discrete time step, mimicking continuous therapy. To simulate individualized treatment response, the dose *d* is treated as a uniform random variable on the interval [0.01, 0.1]. While an alternative way to generate individualized data is to change the parameters associated with tumor growth or radiotherapy response, simply varying the dose proved effective in generating a heterogeneous set of time course data.

Using this approach, we generated 50 “patient” datasets, where the phrase “patient” will refer to simulated patient data from this cellular automaton model throughout this manuscript. The tumor volume predicted by the CA model is scaled to the maximum allowable volume to generate the time course of the relative tumor volume for each simulated patient (Figure 1a). Only the data that represents treatment response over a one month period of time, starting from the dashed line at day 15, is used in our virtual clinical trial. The code used to generate these simulated patients represents a slight modification of the CA code found at https://github.com/storeyk/LVcalibration. This modified code, which details all CA parameter values, is available at https://github.com/jgevertz/VCT.

**Fig. 1.**
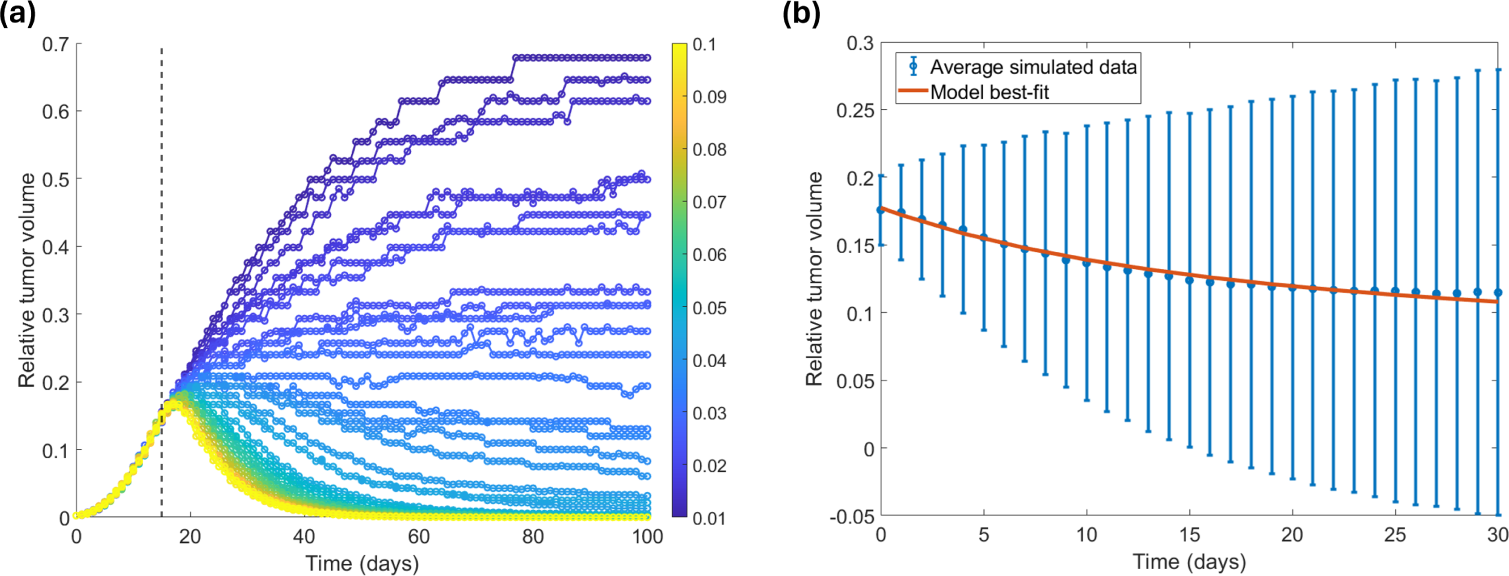
(a) Relative volume of 50 simulated patient tumors generated from the CA model [23] with the dose uniformly distributed on [0.01, 0.1]. The colorbar indicates the dose that was used to generate each simulated dataset. Dashed line indicates onset of treatment. (b) Model best-fit (red) to the average of the truncated tumor volume data (blue).

### 2.2 Toy Model and Fits to Synthetic Patient Data

Our virtual clinical trial next requires a fit-for-purpose and validated mathematical model, as detailed in Steps 1 through 2 of Craig et al. [21]. As our goal in this study is to quantify the impact that the methodology for generating plausible patients has on VCT predictions, we deliberately choose a very simple ODE model of tumor growth and treatment response:

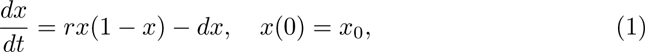

where 0 *≤ x ≤* 1 represents the relative tumor volume, *r* is the logistic growth rate, *d* is the drug-induced kill term, and *x*_0_ the initial relative tumor volume. As this model is an autonomous, one-dimensional ordinary differential equation, its solutions must be monotonic. Further, the model has two steady-states: 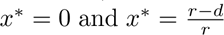, the latter of which is only biologically relevant when *r > d*. Whether or not a solution decreases to the zero or nonzero steady state, or increases to the nonzero steady state, is fully determined by whether *r < d* and whether *x*_0_ is above or below the stable steady-state. Thus, the conclusion of our virtual clinical trial, which is exploring whether a VP has a tumor that grows or shrinks, will be independent of the time horizon for which we solve this differential equation.

To determine if this simplistic model is an adequate representation of the patient data, we use the built-in MATLAB function *fmincon* to minimize the cost function:

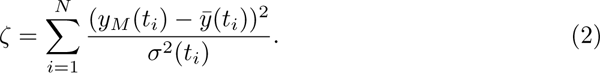

In the cost function, *y_M_* (*t_i_*) represents the model-predicted volume at time *t_i_*, *y*(*t_i_*) represents the average volume in the patient data at time *t_i_*, and *σ*^2^(*t_i_*) represents the variance in the patient data at time *t_i_*. As shown in Figure 1b, this simple ODE model is able to well describe the average of the patient data using parameter values described in Table 1.

**Table 1.** Optimal parametrization of model (1) and how that parametrization was used to determine the features of the normal and uniform prior distributions.

| Parameter | Best Fit | Normal Prior |  | Uniform Prior |  |
| --- | --- | --- | --- | --- | --- |
|  |  | Mean | Std | Min | Max |
| $r$ | 0.3355 | $\mu_r = 0.3355$ | $\sigma_r = 0.75\mu_r$ | $\max\{0, \mu_r - 3\sigma_r\}$ | $\mu_r + 3\sigma_r$ |
| $d$ | 0.3076 | $\mu_d = 0.3076$ | $\sigma_d = 0.75\mu_d$ | $\max\{0, \mu_d - 3\sigma_d\}$ | $\mu_d + 3\sigma_d$ |
| $x_0$ | 0.1775 | Fixed at $x_0 = 0.1775$ | | Fixed at $x_0 = 0.1775$ | |

Given the excellent fit of the ODE model to the average of the patient data (Fig 1b), this ODE model will be used as the underlying mathematical model to describe the patients for our virtual clinical trial. We note that this model does not necessitate any sensitivity or identifiability analysis (Step 3 in [21]), as we are intentionally considering an overly simplistic two-parameter model so that our emphasis can be on VCT design rather than on features of the data or the model. In what follows, we will consider two commonly used methods for generating plausible patient populations that we call the “accept-or-reject” and the “accept-or-perturb” method.

### 2.3 Methods for Generating Virtual Patients

For Step 4 of a VCT [21], we test two inclusion/exclusion criteria that are commonly used for creating plausible patients, each of which requires a prior distribution for each model parameter. The parameters that vary across individuals in this VCT are the intrinsic tumor growth rate *r* and the drug-induced death rate *d*.

In the first method, that we call the “accept-or-reject” method, a model parametrization is determined by randomly sampling (a subset of) parameters from a normal distribution, while rejecting any negative parameters [6, 7]. As the normal distribution is used in all implementations of this method that we are familiar with [6, 7, 21], we will refer to this as the “standard prior” for the accept-or-reject method. A parametrization is considered a plausible patient, and added to the plausible population, if the model-predicted volume trajectory corresponding to the parametrization falls within the defined feasible region *F* (see schematic in Figure 2), where

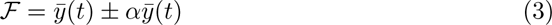

**Fig. 2.**
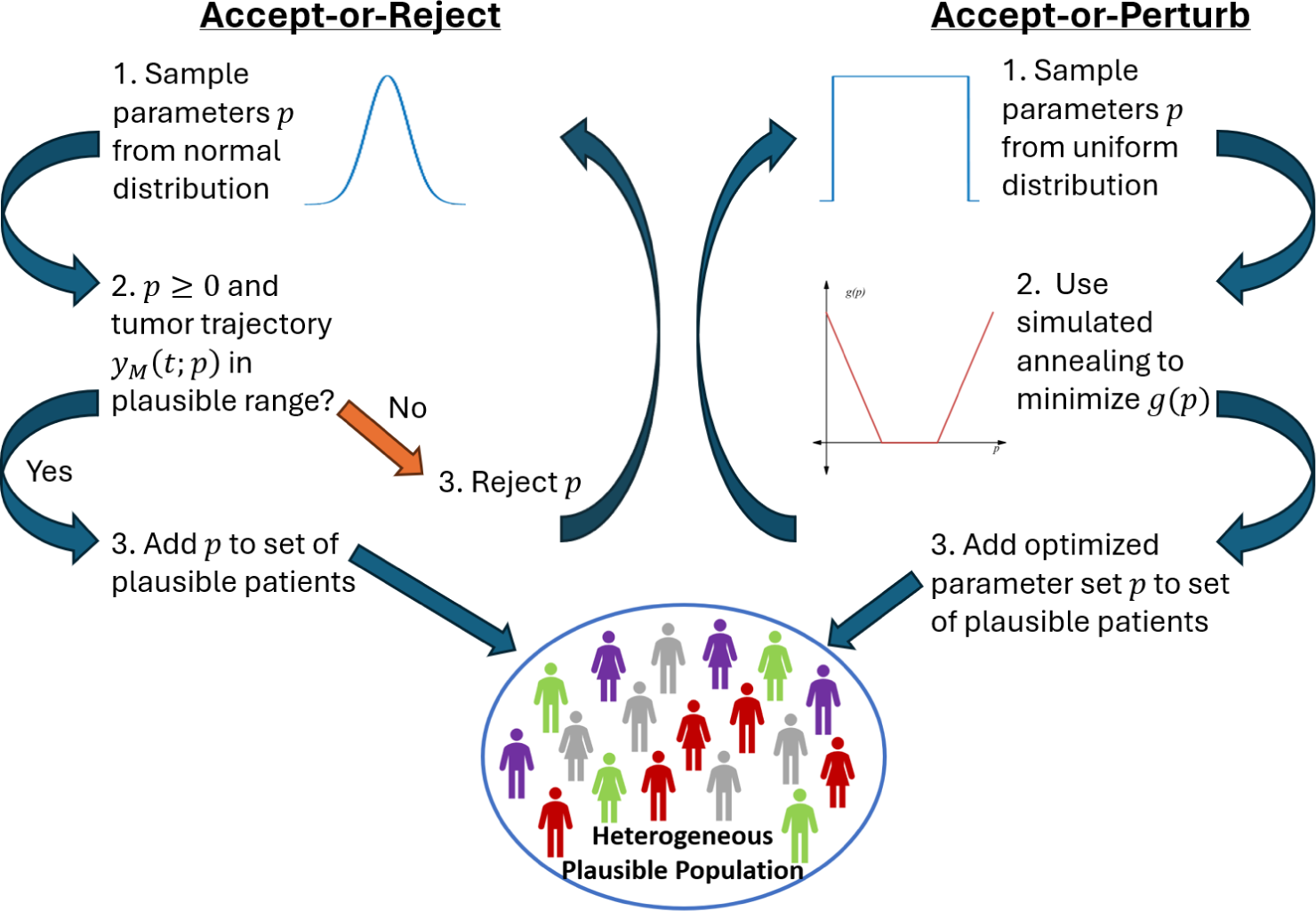
Schematic of the accept-or-reject (left) and the accept-or-perturb (right) method for the generation of plausible populations.

and *α* = 3 unless otherwise stated. In other words, we define the feasible region as being within three standard deviations of the mean patient trajectory *y*(*t*) at each time point *t*. If any portion of the tumor trajectory falls outside *F*, the parametrization is rejected, as visualized in Figure 3a. The accept-or-reject method is an example of Approximate Bayesian Computation [24, 25].

**Fig. 3.**
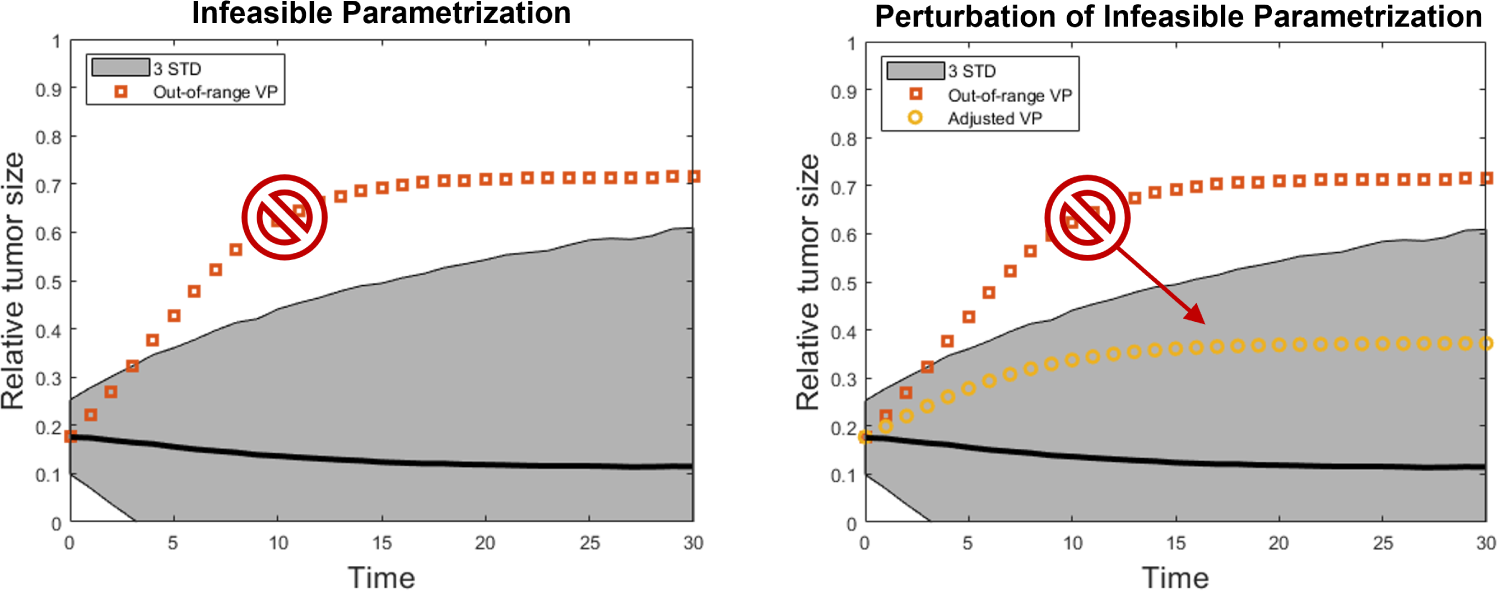
Visualization of how virtual clinical trial methods handle ‘out of region’ parametrizations. The black curve is the mean of the patient data, *y*, and the feasible region *F* is shown in grey. (a) Accept-or-Reject automatically rejects any such non-feasible parametrizations, whereas (b) accept-or-perturb attempts to perturb the parametrization so that the trajectory falls within the feasible region.

In our implementation of the accept-or-reject method, we chose to fix the initial condition at the best-fit value to the average of the patient data (Table 1). The mean of the normal distribution of each variable parameter (*r* and *d*) is set to the best-fit value of the parameter when the model is fit to the average of the patient data (Table 1), and the standard deviation is fixed to be three quarters of the parameter’s mean (*σ_r_* = *βµ_r_, σ_d_* = *βµ_d_* with *β* = 0.75), though, we will also explore the impact of changing the standard deviation later in this study. In our study, we also test a non-standard prior for this method, a uniform prior (details described below in our description of the accept-or-perturb method).

The “accept-or-perturb” method was created by Allen et al. [26] as an approach to generate heterogeneous virtual patients while allowing for an exploration of parametric uncertainty. In this method, a model parametrization is determined by randomly sampling (a subset of the) model parameters from a uniform distribution with user-specified lower and upper bounds. As with the accept-or-reject method, because this method was designed using a uniform distribution [26], we call the uniform distribution the “standard prior” for the accept-or-perturb method. Each parametrization *p* = (*r, d*) is then optimized using simulated annealing (<u>simannealbnd</u> in MATLAB) to ensure that the volume trajectory corresponding to the virtual patient parametrization falls within the feasible region *F*. The objective function to be minimized using simulated annealing is [7, 26]:

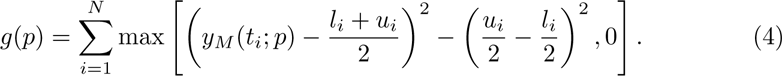

Here, *y_M_* (*t_i_*; *p*) denotes the model output at time *t_i_* for parameter set *p*, and *l_i_* and *u_i_* denote the *i^th^* plausible upper and lower bounds of the tumor volume data at time *t_i_*. By definition, *g*(*p*) assigns a value of 0 (the minimum possible value) only when each time point lies within the feasible region *F*. If a parametrization *p* happens to correspond to *g*(*p*) = 0, then the parametrization is automatically a plausible patient. When a parametrization *p* results in *g*(*p*) *>* 0, simulated annealing is used to perturb *p*. If the optimization converges, the resulting perturbed parameter set then meets the definition of a plausible patient (see schematic in Figure 3b).

In Allen et al. [26], this collection of PPs composes a plausible population. From this PPop, a virtual population is determined using an optimization procedure to find the subset of the PPop that best-matches a set of observed population data according to a measure of statistical similarity [8, 26]. In scenarios where no prior knowledge is available for the final Vpop distribution one simply assumes that the VPop is equivalent to the PPop [8]. Thus the terms plausible patient (PP) and virtual patient (VP) will be treated as equivalent. Similarly, the terms plausible population (PPop) and virtual population (VPop) will also be treated as equivalent. We do note that an advantage of analyzing plausible populations rather than data population-matched VPops is that the PPops do not inherit the demographic biases found in the real-world patient populations [3].

As in our implementation of the “accept-or-reject” method, for the “accept-or-perturb” method we fix the initial condition across virtual patients and allow (*r, d*) to vary across individuals. We again fix the standard deviation of each prior to be three quarters of the parameter’s mean. The uniform distribution is then defined such that the minimum value is set to be *α* = 3 standard deviations below the mean (or 0 if this value is negative), and such that the maximum value is *α* = 3 standard deviations above the mean (Table 1). In our study, we also test a non-standard prior for this method, a normal prior (details described above in our description of the accept-or-reject method).

In summary, we will be testing two parameter priors (uniform and normal, which was also done in Kolesova et al. [22]) and two inclusion/exclusion criteria (accept-or-reject and accept-or-perturb) for plausible patients. We will consider all possible combinations of these choices, thus allowing us to home in on the impact that both the prior and the inclusion/exclusion criteria have on the composition of the virtual population and the predictions of the virtual clinical trial. We will also study how parameters associated with VCT design (spread of the prior as determined by *β*, extent of the feasible region *F*, and the number of virtual patients) impact VCT outcomes.

## 3 Results

Herein, we quantify how the design of a virtual clinical trial impacts the variability in the VPop and the predictions about treatment response for our example trial. Working with the proposed toy model in the context of synthetic data allows for a more robust study of the impact of the parameter prior distributions, and the method used to create the posterior distributions (which can be thought of as our statistical description of the VPop).

### 3.1 Impact of VCT Prior Distribution

We first compare virtual clinical trial results when a single method (accept-or-reject or accept-and-perturb) is implemented with different prior distributions: a uniform distribution (*u*) or a normal distribution (*n*), whose parameters are both described in Table 1 unless otherwise specified. Because of the shape of these distributions, the normal prior is biased towards values closer to the mean. The uniform distribution, on the other hand, allows for more parameter values farther from the mean (particularly, values larger than the mean, given the truncation at zero) to be considered with equal probability to those values closer to the mean. As a result, the normal distribution is more likely to generate plausible model parameterizations. In particular, we find that implementing the accept-or-reject method on model (1) resulted in 302 random samples with non-negative parameters being rejected in order to generate 1000 virtual patients. This equates to about 76.8% of random parameterizations being accepted. However, when a uniform distribution was used, only approximately 69.8% of random (non-negative) parameterizations were accepted. Similarly, with the accept-or-perturb method, 75.2% of samples were accepted without perturbation when using a normal distribution, as compared to 68.7% when using a uniform distribution.

The impact of using these priors to generate 1000 VPs, each defined as a pair of *r* and *d* values, with the accept-or-reject method is visualized in the center row of Figure 4. Consistent with the shape of the prior distributions, the normal prior creates a VPop in which the posterior distribution is biased towards the best-fit value of each parameter for the average patient. On the other hand, the uniform distribution results in posterior distributions with larger spreads. This equates to more variability among the patients in the VPop. Interestingly, for the death parameter *d*, accept-or-reject with a uniform prior results in a posterior distribution skewed significantly further right than the prior distribution. This demonstrates that the choice of prior has a significant impact on the *composition of the virtual patient population*.

**Fig. 4.**
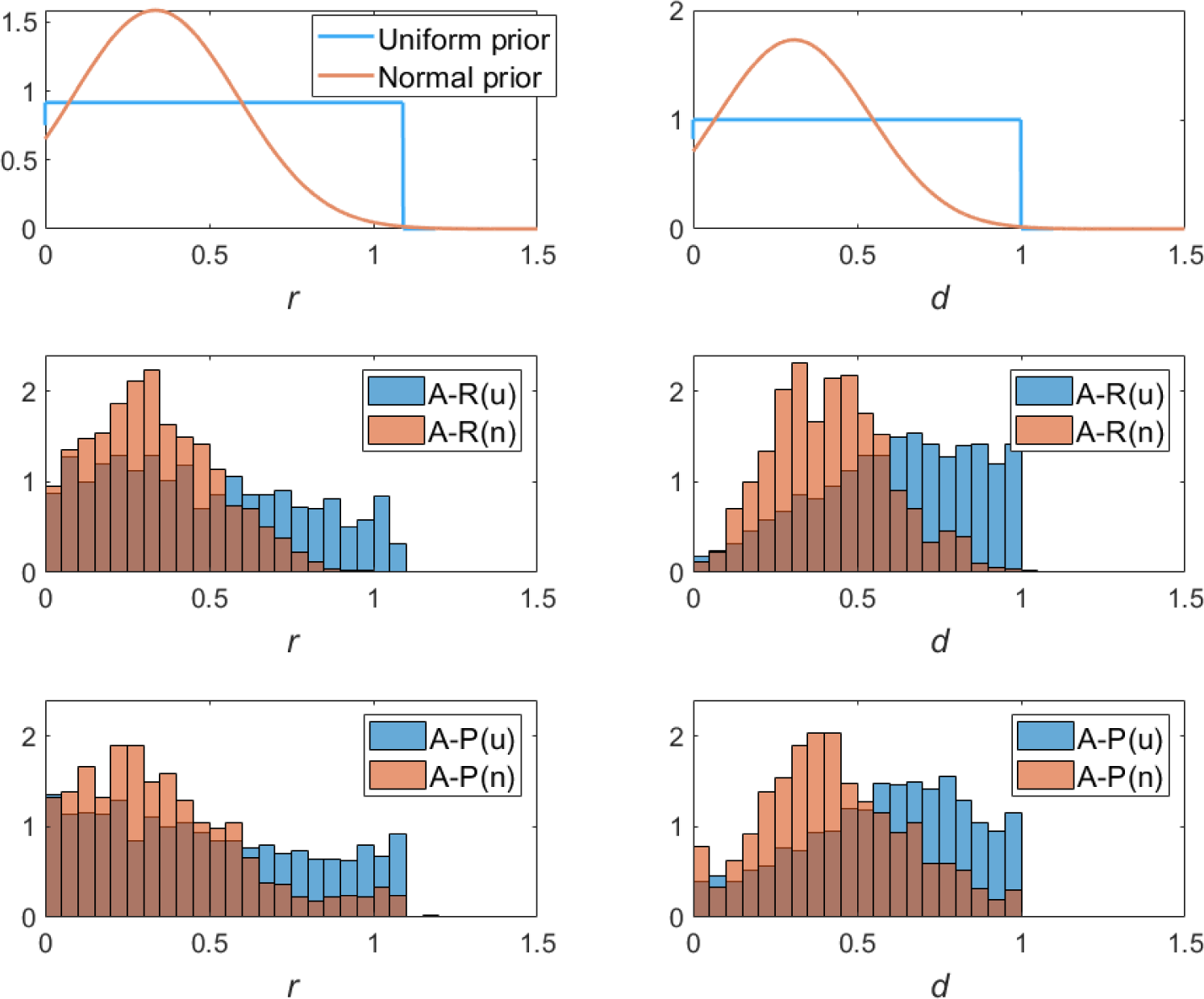
Quantification of the impact of the prior distribution for the accept-or-reject (A-R) method (center row) and accept-or-perturb (A-P) method (bottom row). Results related to uniform distribution are shown in blue and those related to the normal distribution are shown in red. The overlap between the two distributions appears in a deeper red/brown shade. The prior and posterior for the tumor growth rate *r* are shown on the left, whereas the corresponding distributions for the death rate *d* are shown on the right.

It is certainly possible that the bias that emerges based on the prior using accept-or-reject will not emerge using accept-or-perturb. This is because the accept-or-perturb method does not outright reject nonviable parametrizations; instead, it perturbs the parametrization until the corresponding tumor volume trajectory falls within the feasible region *F*. The impact of using a uniform and a normal prior distribution to generate 1000 VPs with the accept-or-perturb method is visualized in the bottom row of Figure 4. Interestingly, the bias of the posterior distribution towards the shape of the prior is evident even using this methodology. Particularly, the posterior of both *r* and *d* are biased towards the mean of the normal distribution when a normal prior was used, whereas the posteriors exhibit significantly more spread when a uniform prior is utilized. This further demonstrates that the choice of prior has a significant impact on the composition of the virtual patient population, even when the VCT method is designed to perturb, rather than automatically reject, non-feasible parametrizations.

### 3.2 Impact of VCT Selection Method

Now we shift our focus from the impact of the prior distribution to the impact of the method and criteria used to exclude a parametrization from the VPop. In the top row of Figure 5, we consider the case of a uniform prior and compare the posterior distributions that result from using accept-or-reject or accept-or-perturb to create a VPop consisting of 1000 VPs. The analysis is repeated for a normal prior in Figure 5 (middle row). Interestingly, in both cases, we find that the choice of accept-or-reject versus accept-or-perturb has minimal impact on the heterogeneity of the VPop, as visualized through the significant overlap in the posterior distributions in Figure 5 (left, top and middle). Though we do observe more parametrizations with large *r* values in the posterior distribution for the accept-or-perturb method. This is especially noticeable in the top right section of the scatter plot (Figure 5 middle right), which uses a normal prior. This is attributable to the fact that the perturbation process makes it easier to reach extremal areas of parameter space.

**Fig. 5.**
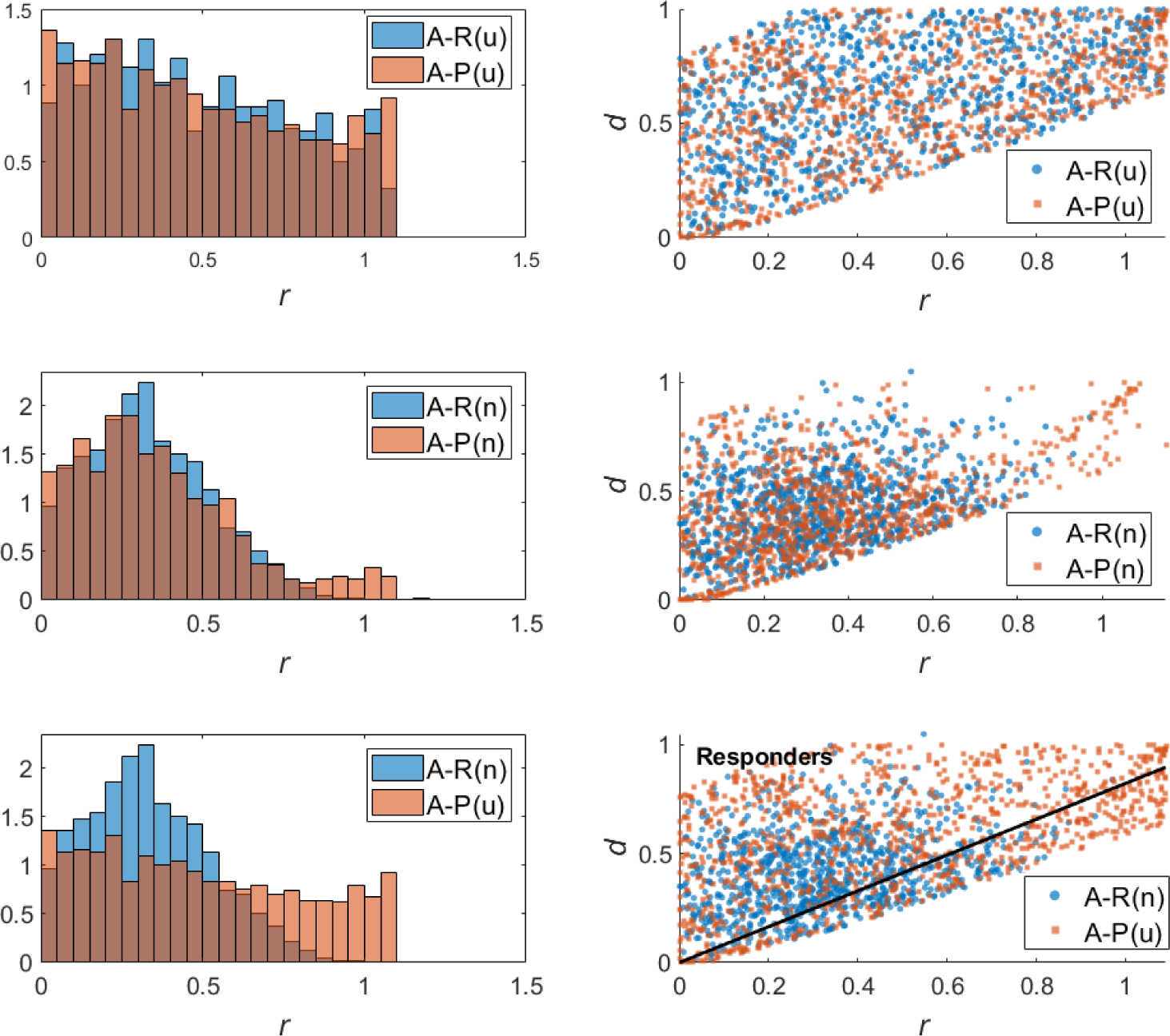
Quantification of the impact of the inclusion/exclusion criteria using a uniform prior distribution (top) or normal distribution (middle). Accept-or-reject (A-R) results are shown in blue and accept-or-perturb (A-P) results are shown in red. The bottom row shows posterior distributions for the standard pairing of accept-or-reject with a normal prior and accept-or-perturb with a uniform prior. The solid black line, defined in eqn. (5), separates responders from non-responders in model (1).

It is of note that the standard “pairing” of prior distribution and method/selection criteria is to use a normal prior for accept-or-reject and a uniform prior for accept-or-perturb. A comparison of the heterogeneity of the VPops generated via these standard methods is shown in the bottom row of Figure 5. The parametrizations of the VPs generated by accept-or-perturb are scattered over a larger region of the *r−d* plane than those generated by accept-or-reject. Based on the above analyses, the fact that accept-or-perturb creates a more heterogeneous virtual population is largely attributable to the use of the uniform prior, and not the selection criteria.

### 3.3 Impact on VCT Prediction

Next we compare how the design choices impact the prediction made by the VCT. We define the “outcome” of our virtual clinical trial as the percent of responders in the VPop, where a responder is any VP for which the corresponding tumor (as determined by model (1)) decreased in volume over the time course of the study. It is of note that in the simulated patient data (Figure 3a), 74% of the patients are classified as responders. As with all subsequent analyses, prior distributions are defined as in Table 1, unless otherwise specified. 1000 virtual patients were generated per method, using the feasible region *F* defined in eqn. (3) with *α* = 3 unless otherwise specified.

Despite the fact that the prior had the larger influence on the posterior distribution, we surprisingly observe that the selection method has a larger influence on the outcome of the trial. As shown in Figure 6, accept-or-perturb predicts fewer responders (69.7% for uniform prior and 67.6% for normal prior) in the trial than accept-or-reject (78.1% for uniform prior and 74.0% for normal prior). As the actual response rate in the patient population is 74%, this means that accept-or-reject is equaling or over-estimating the true response rate, whereas accept-or-perturb is underestimating this rate.

**Fig. 6.**
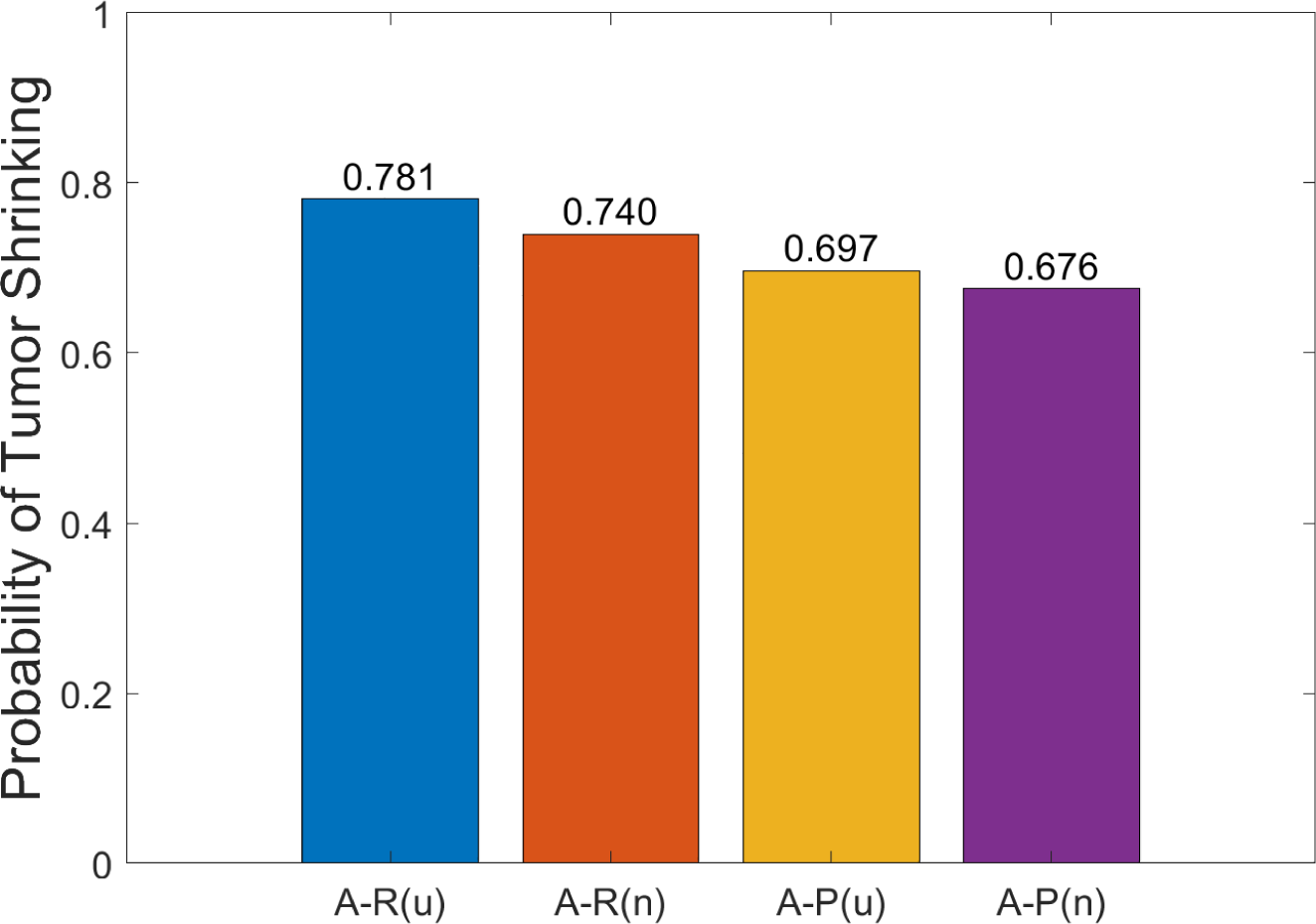
Fraction of responders in a VPop generated using accept-or-reject (A-R) or accept-or-perturb (A-P) using either a uniform (*u*) or a normal (*n*) prior distribution.

To understand why accept-or-reject consistently predicts a larger response rate than accept-or-perturb, in the bottom right of Figure 5 we visualize the line in parameter space that separates responders from non-responders in model (1) when *x*_0_ = 0.1775. This separating line was calculated using the fact that non-responders in (1) must have a positive nonzero steady-state 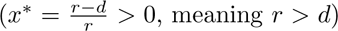 and start below this steady state, giving rise to the constraint that:

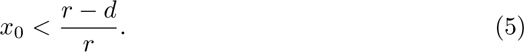

Comparing this boundary to the distribution of points in parameter space shows that there are significantly more red squares (associated with accept-or-perturb) than blue circles (associated with accept-or-reject) in the non-responder region of parameter space. Thus the response rate is lower using accept-or-perturb. We speculate that this occurs because accept-or-perturb can “reach” regions of parameter space that are not likely to be reached by the accept-or-reject method (see Figure 4).

This trend proved to be quite robust across virtual clinical trial parameters. In Figure 7a we explore the impact of changing the standard deviation on the prior distributions. With the exception of using the smallest tested standard deviation of *β* = 0.25 (meaning the standard deviation is a quarter of the mean of the parameter), accept-or-perturb consistently predicts less treatment responders than accept-or-reject. The fact that *β* = 0.25 is an outlier to the trend is not surprising, as using a standard deviation that is too small greatly limits the range of allowable parameters which over constrains the behavior of plausible patients. As stabilization of the plots in Figure 7a indicate, it is desirable to be “greedy” on the prior. The methods are already designed to reject or perturb nonviable parametrizations, and the only cost of a prior with a large standard deviation is a computational cost - it takes longer to build a virtual population of a fixed sample size when more parameters are rejected or perturbed. However, this extra computational cost results in more heterogeneous plausible patients.

**Fig. 7.**
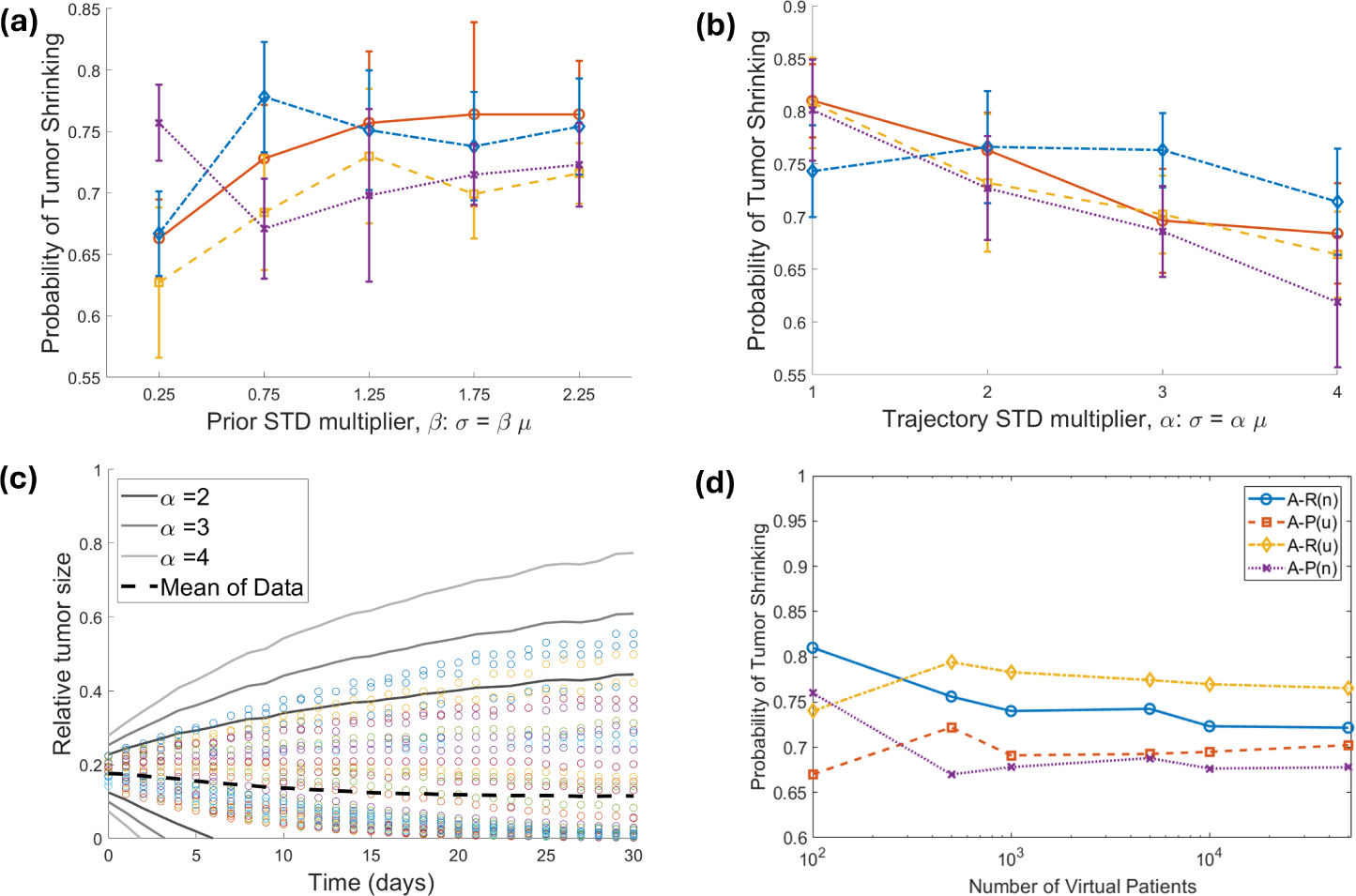
Impact of varying VCT parameters. (a) Predicted probability of treatment responders as a function of the *β*, the standard deviation multiplier of the prior distribution. *N* is fixed at 100 and the experiment is repeated 10 times. Standard deviation multiplier of feasible region *F* is fixed at default value of *α* = 3. (b) Predicted probability of treatment responders as a function of *α*. *N* is fixed at 100, *β* is fixed at its default value of 0.75, and the experiment is repeated 10 times. (c) Visualization of patient data, and acceptance region for the various values of *α* studied in (b). (d) Predicted probability of treatment responders as a function of the number of virtual patients included in the VPop with all other parameters set at default value (*α* = 3, *β* = 0.75).

In Figure 7b, we quantify the impact of the size of the feasible region 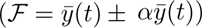 by allowing *α* to vary from 1 to 4. Besides the case of *α* = 1, we still observe that accept-or-perturb predicts fewer responders than accept-or-reject. In the case of *α* = 1, the acceptance region is so restrictive that a large fraction of actual patient trajectories would be excluded from the virtual population. In fact, even the case of *α* = 2 restricts a number of actual patients from being included in the VPop (Figure 7c). Figure 7b demonstrates that *α* = 3 is a “sweet spot” for the definition of the feasible region: it is large enough to include all simulated patients, while also leaving room for some outlier trajectories. It is not so large, however (as in the *α* = 4 case) that trajectories that differ significantly from what is observed in the patient population would be included in the VPop. Increasing the size of the acceptance region generally results in a decrease in the predicted fraction of responders, though the impact (and the deviation from the 74% response rate found in the patient data) is more substantial when using accept-or-perturb.

Finally, in Figure 7d we explore the impact of the size of the VPop on treatment response. We find that predictions for the fraction of responders for all four prior/inclusion criteria combinations begin to stabilize at a sample of *N* = 1000. Below this value, we find that the fraction of responders increases as a function of population size for VCTs using a uniform prior, but decreases as a function of sample size for VCTs using a normal prior. That said, we do not hypothesize that there is anything biologically significant about these particular trends. Instead, this is likely an artifact of not having a sufficiently large sample size. The general trend that accept-or-perturb predicts fewer responders than accept-or-reject holds for sample sizes of *N* = 500 and larger. Though, we do note that the predictions stabilize quicker for VCTs that used the uniform prior. This is likely because the normal prior is less likely to sample extreme parameter values, and thus requires significantly more samplings to include outlier VPs that the uniform prior could more readily sample.

## 4 Conclusion

Various methods for creating plausible populations for virtual clinical trials have been proposed in the literature. In this work, we assess how the design choices for creating plausible patients impacts the heterogeneity of the plausible population and the predictions of the VCT. To isolate the impact of VCT design choices, we work with simulated patient data and a simple, toy model of tumor growth in response to the treatment. In this controlled setting, we study the impact of the following VCT design choices: the prior parameter distribution (either uniform or normal of various standard deviations) and the method for selecting parametrizations for a plausible population (either accept-or-reject or accept-or-perturb, with various definitions of the feasible tumor trajectory).

Conducting a trial using each of the inclusion/exclusion methods with the two prior distributions revealed that the prior distribution has the most significant impact on the heterogeneity of patients in the plausible population. In particular, a uniform prior resulted in posterior distributions with greater spread than is achieved using a normal prior. When a normal prior is used, accept-or-perturb does result in a posterior with greater spread than accept-or-reject does. We next used the resulting posterior distribution as the plausible population for our VCT. Surprisingly, the inclusion/exclusion criteria had a larger impact on trial outcomes (defined as percent of responders) than the choice of prior distribution. We particularly found that the PPops created using the accept-or-reject method had a higher percentage of responders than those created with the accept-or-perturb method. In each case, using a uniform distribution also resulted in a higher percentage of responders. We found that, except in extreme cases, the response probability was robust to the following VCT design choices: prior standard deviation, extent of feasible region for tumor trajectories, and number of plausible patients in the PPop. This is in contrast to the sensitivity observed to the choice of inclusion method (and less so, the prior).

The power of virtual clinical trials lies in their ability to quantify the impact of heterogeneity on treatment response. However, independent of the methodology used to generate plausible patients, there are a number of shortcomings to be aware of. The results of a VCT are only as good as the model itself [6]. Hence, the process of model validation is an essential step [21, 27] before conducting a VCT. Another consideration is that there is no rigorous way to define the feasible region for a virtual patient trajectory. This has the potential to introduce bias into the VCT based on which parametrizations get included in the VP cohort [6].

Yet another complication is that while a PPop can help explore the broad range of responses the model can produce, the outcome may not necessarily reflect the distribution of population-level data; that is, the probability of observing each outcome [8]. In this work, and in other scenarios where such population-level data is not available, one can follow the lead of Surendran et al. [7] and treat the plausible population as the virtual population. Even when population-level data is available, a downside of “matching” the VPop to population-data is that the VPop will then recreate the biases inherent in that dataset [3].

In the case where such data is available and expected to be an unbiased representation of the patient population, a number of approaches have been proposed to identify a virtual population from a plausible population. Klinke proposed a method for weighting each plausible patient to ensure that the descriptive statistics of the VPop are optimally matched to the population-level statistics [18]. The computationally-intensive nature of this approach has resulted in others innovating on this idea. In the Mechanistic Axes Population Ensemble Linkage (MAPEL) algorithm, weights are assigned to “mechanistic axes” rather than to individual plausible patients [19]. The defining of the mechanistic axes, which can be thought of as groupings of parameters/features, complicates the use of this method, however. In Allen et al. [26], a methodology is introduced for identifying a VPop from a PPop that calculates a probability of inclusion for each plausible patient and uses those probabilities to select a subset of the PPop that best matches the descriptive statistics of the population-level data. Other methods for generating virtual populations have also been proposed [9, 22, 28].

We intentionally conducted this analysis in a controlled setting using simulated patient data and a two-parameter toy model to isolate features of VCT design that influence trial outcomes. However, it is important to note that both accept-or-reject and accept-or-perturb can be inefficient on more complex mathematical models both because these are more time-intensive to solve than simpler models, and because the majority of parametric samplings will not result in an (initially) plausible patient [29]. These more complex models could be mechanistic-based ODEs like seen in quantitative systems pharmacology [6], but also partial differential equations [30] and even agent-based models [31].

For these more complex models, the methods used herein necessitate either generating and testing many more parametrizations than desired in the VPop (if using accept-or-reject) or many executions of simulated annealing (if using accept-or-perturb). To generate VPops for these higher dimensional models, alternative methods have been proposed to speed up the process of generating a plausible patient. For instance, Meyers et al. show how a surrogate machine learning model can be trained on the full mathematical model and used to rapidly pre-screen for parametrizations that result in plausible patients [29]. Though, this and other surrogate modeling approaches still must contend with the computational time to develop the surrogate model, not to mention possible inaccuracies in the surrogate model’s representation of the true model [32]. In Derippe et al. [32], an approach is developed to improve the computational efficiency of VP development (particularly the acceptance/rejectance step) under the assumption of monotonicity of a subset of model parameters with respect to the model output. Beyond the computation costs of working with more complex models, they also generate large amounts of data that can be quite challenging to rigorously analyze. Machine learning approaches have been proposed to efficiently analyze Vpop behavior [33].

Just as the complexity of the model poses challenges for the development of Vpops, so does the complexity of the experimental data. In these larger, more complicated models, it is typically necessary to run more simulations to appropriately calibrate plausible patients to all available pharmacodynamic biomarkers and endpoints [6]. Further, in the case of multi-modal data, challenges arise in how to define the inclusion criteria for a plausible patient [6]. Another complication that arises in the use of multi-modal data for complex models is that the assumption of parameter independence is less likely to be valid. Methods have been developed that move beyond the assumption of a univariate distribution for each personalized model parameter. For instance, Parikh and colleagues have introduced a generative adversarial network architecture to infer joint densities of model parameters [34].

Despite these challenges, we sought to conduct a controlled study to gain insight into how different VCT design choices impact the heterogeneity of plausible populations and the outcomes of the VCT. While we do not believe this work can suggest a single “best” approach for all VCTs, just as argued in [8], it does yield some observations which can guide design choices for future VCTs, particularly in the absence of a large and unbiased population-level dataset to constrain plausible populations. If the goal is to obtain the largest coverage possible of plausible parameter space, we recommend the use of a uniform prior distribution rather than a normal prior (Figure 6). However, the use of a uniform prior does result in a scenario where parameter values far from the mean get represented with a significant probability in the plausible population. If we want to be able to reach these outlier parameter values, but create plausible populations where the parameters are more biased towards the mean, the accept-or-perturb method with a normal prior accomplishes this goal. It is our hope that this foundational understanding of the role of virtual clinical trial design will inform the development of future VCTs that use more complex models and real data.

## Data Availability

All data produced are available online at https://github.com/jgevertz/VCT

## Acknowledgements

The authors thank Katie Storey for her support in implementing the cellular automaton model to generate simulated patient data.

## Declarations

The authors have no relevant financial or non-financial interests to disclose. The code used to generate all the data presented in this manuscript is available at https://github.com/jgevertz/VCT.

